# Population mobility and migration in health policies: policy as discourse analysis of Zambia’s Neglected Tropical Disease Masterplans 2011-2022

**DOI:** 10.1101/2025.08.01.25332586

**Authors:** Patricia Maritim, Margarate Nzala Munakaampe, Joseph Mumba Zulu

## Abstract

**Background:** Despite population mobility being on the rise globally, it remains neglected in the development of national health policies. This study sought to examine how population mobility is problematized in National Neglected Tropical Disease Masterplans and how these representations shape policy implementation within 4 NTD endemic districts in Zambia.

**Methods:** We used Policy as discourse analysis by guided by Bacchi’s “What’s the Problem Represented to be?” (WPR) approach. We reviewed all four NTD Masterplans that have been developed in Zambia between 2011 and 2022. Interviews on everyday implementation experiences were conducted with key policy actors (n=20), community health workers (n-34) and community members (n=35) guided by the Consolidated Framework for Implementation Research. Data was analysed using critical discourse analysis informed by the WPR approach.

**Results:** Across all four Masterplans, the construction of migration and its role in NTD prevention, control and treatment was scant indicating limited recognition of the role that migration plays on health. Migrant groups recognized were forced migrant (refugees) or voluntary migrants (foreign tourists), labour migrants (fishing or farming) or environmental migrants.

Assumptions underlying these constructions of migration include territoriality and associated internalization practices, medical nativism, migrant deservingness and universalization. Silences in the Masterplans included failure to account for local migration patterns, lack of migrant sensitive services and lack of clarity on entitlement of health services.

**Conclusions:** Though the mobility is poorly problematized in the Masterplans, through policy design and implementation can strengthened such as using more comprehensive and standardized definitions of migration and migrant groups in public policies, capturing migration data and using it to inform decision making as well as establishing accountability mechanisms for policy actions, training of frontline workers using standards and engagement of migrant and mobile populations in migration governance.

**AUTHOR SUMMARY:** Zambia has a robust migration governance framework aimed at enhancing population mobility. In order to identify if the policy intent of these governance frameworks in bringing about orderly, safe regular and responsible migration and mobility of people was being translated into health, we examined the problem representations of population mobility in National NTD Masterplans. We found that across all four Masterplans, the construction of migration and its role in NTD prevention, control and treatment was scant indicating limited recognition of the role that migration plays on health. There was an absence of data on the magnitude of migration related inequalities and how migration interacts with other dimensions of social identity. Key assumptions underlying these representations of mobility include territoriality where the Masterplans view borders as fixed orderings of space with cross border migration being seen as responsible for new diseases that are non-endemic in Zambia (onchocerciasis) or reintroduction of NTDs which have been eliminated (leprosy). The failure of the Masterplans to establish strong links between migration and NTD prevention, control, and treatment could be linked to a broader policy agenda surrounding migration nationally, regionally and internationally.

## INTRODUCTION

Population mobility is a defining feature of the world we live in, as people move for a variety of reasons such as climate change, political conflict, forced displacement, employment or education (1). Migration involves a complex set of dynamic processes associated with human mobility that captures two critical aspects; spatial distance (where people go) and temporal duration (how long they stay) (2,3). Therefore, a holistic view of this complexity means a recognition of a broad range of forms of entry and legal status (permanent legal residents, temporary work visa, undocumented) as well as temporal mobility patterns (seasonal movement vs continued residence in a single location) (4). Common categories of mobile populations include international (cross border) migrants, internal migrants, labour migrants and forced migrants such as refugees, human trafficking and climate change refugees (3,5). Currently, it is estimated that there are over 281 million international migrants and more than 750 million internal migrants globally, of whom 62.5 million are internally displaced due to conflict and violence (6). The African region hosts 25.1 international migrants with most African migrants moving within the continent (7).

Over the recent past, there has been increased attention to promoting the physical and mental health of migrants (8) which is evident from the adoption of different international commitments such as World Health Assembly Resolution 61.17 on the Health of Migrants, United Nations Global Compact for Safe, Regular and Orderly Migration, the UN Global Compact on Refugees and WHO’s Global Action Plan for Promoting the Health of Refugees and Migrants 2019-2030. Migrant health has also risen as a global health research interest with the WHO listing the “identification of effective and sustainable models of health care for migrants, refugees and other displaced persons in humanitarian settings in LMICs and fragile settings” as one of its global research priorities (9). This is particularly important because migrant populations experience health inequities defined as preventable differences in health outcomes and inequalities in access to healthcare and the outcomes of healthcare between population groups who occupy different positions in social, economic and political structures (10–12). These inequities arise from exposure to spaces of vulnerability throughout migration journeys that contain conditions which increase their likelihood of exposure to and acquisition of diseases (13). Such spaces are made possible through drivers such as conflict trauma, violence and other human rights violations, exclusionary migration policies, unsafe inadequate shelter, food and housing; racism, discrimination and stigma, labour market exclusion and barriers to healthcare and other services (14).

Migrant health is essential in the realization of Universal Health Coverage under Goal 3.8 of the Sustainable Development Goals and Reducing inequalities under Goal 10.7 which calls for countries to facilitate “orderly, safe regular and responsible migration and mobility of people including through the implementation of planned and well managed migration policies” (15). With countries being urged to incorporate population mobility as they develop public health policies and strategies so that they can recognize different types of migrants including refuges, displaced persons and other mobile populations to ensure health systems are responsive to migration (16). Migration aware public policies in particular have been identified as being critical to the reduction of migration related health inequities (5). As they are part of whole system approaches which embed population movement/mobility a central concern during their design and implementation (5,13). The focus on human mobility espoused in migration aware policies is rooted in three main arguments. Normatively, it eliminates the cognitive framing that *others* migrants in relation to the general populations resulting in marginalization (2). On an empirical level, it moves away from reducing migrants experiences with a bias to settlement in destination areas which diminishes the complexity of processes underlying human mobility (2). Finally, it allows us to examine different aspects of mobility that are understudied and whose appreciation can result in the generation of migrant sensitive interventions (2). Vearey has provided examples of strategies that are essential components of a migrant aware policy response include; training of health providers at all levels on migration, mobility and health, recognition of internal mobility within countries, implementing legislation that recognizes the right to health for all people including migrants, having monitoring systems at facility level that ensure that this legislation is being implemented and working collaboratively with other countries for strengthened regional responses (13). Yet despite the need to ensure integration of migration into how health policies are planned, designed, implemented and assessed (17), population mobility remains neglected in the development of national and global health policies (16). In instances where policies include mobility, lack of comprehensive conceptualization could point to migration not being considered a policy priority (18). All of which results in the persistence of migration related health inequities (5,19).

Population movements at local, rural-urban, regional and global levels are responsible for the expansion of the geographical distribution of Neglected Tropical Diseases infection (20) and migrant populations have been identified as being persons never treated for NTD interventions such as mass drug administration and case management for more chronic manifestations of these diseases (21,22). In many endemic countries, national programmes have been developing and using Neglected Tropical Disease Masterplans. In the African region for instance, the WHO Regional Office for Africa through the Expanded Special Project for the Elimination of Neglected Tropical diseases (ESPEN) has been providing technical support to endemic countries to develop and implement National Master plans (23,24). The Masterplans can be thought of as Big P policies which provide a “formal implementation blueprint that maps outs goals, strategies, scope of the intervention, timelines and milestones as well as other metrics of disease elimination programme success” (25). Masterplans provide a strategic approach to the implementation of evidence based interventions that either prevent or treat disease manifestations (preventive chemotherapy or intensified disease management) or address the greater social determinants of health that facilitate their transmission and severity (water and sanitation practices, vector control and veterinary public health measures)(26). They help ensure a better fit of NTD interventions into the contexts they are being implemented or provision of support to organizational contexts to better support the implementation of these interventions (27). They accomplish this by promoting integrated programming, leveraging existing resources and expertise of different partners and facilitating linkages across different sectors (28).

If NTD Masterplans are to address health inequities, including those faced by migrant groups, they ought to account for the complex socio-cultural factors that influence patient’s experiences ranging from disease susceptibility to living with life altering morbidity (29). Aligning the policy intent of migration aware health policies with how they are designed and implemented is essential to mitigate against repeatedly leaving the migrants out of interventions (30). However, policy content is often formulated within “pre-set political parameters” that are emblematic of a broader policy agenda surrounding migration nationally, regionally and internationally (31). Further, policymaking processes are not neutral and are influenced by the interests, values and normative assumptions of different policy actors (32). Identifying the gaps between policy rhetoric and implementation practicalities can reveal the underlying policy paradigms and their roles in upholding social conditions that include or exclude population groups such as migrant populations is therefore necessary (33,34). We sought to examine how population mobility is problematized in National NTD Masterplans and how these representations shape policy implementation within 4 NTD endemic districts in Zambia. Specifically, we were intent on examining the adequacy of policy’s conceptualization of migration and if its underlying assumptions i.e. level of migration awareness and how this informs the selection of strategies (35). Further, we wanted to understand how the strategies outlined in the policy privilege different social groups over others and why.

## METHODS

### Study setting

Zambia has had a long history of welcoming refugees and asylum seekers with an open door policy (36). It is a signatory of a variety of regional and international instruments that are critical to the promotion of the right to health for migrant and mobile populations such as Convention Relating to the status of Refugees, Global Compact to Safe, Orderly and Regular Migration (36). Further, it has formulated and enacted different laws and policies for effective migration governance including the Immigration and Deportation Act of 2010 and National Migration Policy. International migrants make up 0.8% of the Zambian population and are largely from neighbouring countries such as the Democratic Republic of Congo and Angola (37,38). The main reasons for immigration into the country has been seeking asylum or refugee status, following family as well as for economic reasons (37,38). Internal migration is estimated to be at 16.8% (37).

Zambia’s health system is categorized as being migrant friendly where: migrant groups are conceived as homogenous group, strategies are targeted towards individual level rather than system level responses and population mobility is largely centered on cross border mobility (15). Cross border collaborations and multisectoral collaborations are proposed as some of the main strategies to address population mobility. Migrants face challenges navigating local facilities due to language and communication barriers resulting from the absence of professional interpretation services and culturally incongruent services (21,39) as well as limited information on how to access health services especially among young migrants (36). Refugees in settlement camps access healthcare through in camp health centres. However limited infrastructure to accommodate health providers means that they are not always available on site (40). Most of these health centres also operate without adequate equipment, supplies and essential medicine which means that they cannot handle complex cases. Poor road infrastructure also means that they face shortages and have poor ambulatory services due to frequent breakdowns (40).

### Study design

We applied a post structuralist inquiry to deconstruct the content of the Masterplans, analysed as political constructs which had evolved over time in response to changes in cultural representations, socio-economic processes, social practices (41). We used Bacchi’s post structuralist policy analysis technique, Policy as discourse analysis, to assess how social processes and interactions influence policy formulation, where policy actors not only respond to societal problems but create them in the policy proposals recommended as solutions (42,43). Our conceptualization of policy was that of an “unstable compromise” made by state and non-state actors with differing interests and power which may result in strategies that may either reduce, mitigate or exacerbate health inequities (44). We were guided by Bacchi’s “What’s the Problem Represented to be?” the (WPR) approach that uses a series of questions to conduct policy are discourse analysis (43). In our case to understand how NTD Masterplans have constructed migration and human mobility as policy problems in discourses based on the proposals put forth to address them (42,45). Through the use of qualitative methods, we used this approach to illustrate how “reality comes to being” (42). We used 4 WPR questions including one question that sought to examine equity objectives more broadly (46).

1. What is the problem of health equity represented to be in NTD Masterplans 2011-2026?
2. What is the problem of human mobility and migration represented to be in NTD Masterplans 2011-2026?
3. What assumptions underlie these representations of human mobility?
4. What is left unproblematic in current representations of human mobility? Where are the silences?

### Data collection

#### Document reviews

We reviewed all NTD Masterplans that have been developed in Zambia. The country has had four NTD Masterplans developed and reviewed in alignment to the WHO Global NTD Roadmaps; 2011-2015, 2015-2020, 2019-2023 and most recently 2022-2026 (47). The scale up of interventions in endemic districts across the country guided by the Masterplans, over the past ten years, has led to significant progress in the realization of disease elimination targets such as reduction in the number of individuals requiring interventions against lymphatic filariasis and trachoma (48–50). Nevertheless, disease elimination targets initially set for 2020 (51) have yet to be realized particularly within hardly reached populations such as migrant groups (52).

#### Interviews

Qualitative data collected through in-depth interviews was intended to understand how different actors within communities where Masterplans are implemented including clients, families and implementers experience the policies in their everyday lives within their regular environments (33,53). This would show how these problem representations of human mobility in the NTD masterplans play out in real world settings (43). Data was collected in 4 purposively sampled districts across 4 provinces; Livingstone, Nkeyema, Kapiri and Luangwa Districts. These districts are endemic to the four major NTDs in Zambia; trachoma, lymphatic filariasis, schistosomiasis and soil transmitted helminths (10), have been implementing different interventions outlined in the NTD masterplans and had different human mobility patterns (1).

A total of 20 Key informant interviews were conducted with different stakeholders at provincial and district levels. Stakeholders were drawn from different institutions involved in the implementation of one of the five prevention and control strategies under the National NTD Masterplan. To be eligible to participate, the organizations must have been actively involved in the implementation of at least one strategy in collaboration with the Ministry of Health i.e. preventive chemotherapy, intensified disease management, water and sanitation practices, vector control and veterinary public health measures (26). Interviews were also conducted from community health workers (=34) who are involved in the implementation of various activities such as mass drug administration, case identification and promotion of environmental sanitation and hygiene. Community members were also invited to take part in the study (n=35). When sampling the community members, research teams included patients who had chronic manifestations of some of the NTDs such as lymphoedema and trichiasis so that patients’ perspectives would also be represented within the study. Table 4 provides a summary of the study participants. Since our main focus was on understanding the context of every day implementation, data collection guides were based on a modified version of the Consolidated Framework for Implementation Research adapted for health equity and for use in low and middle income countries (54,55). All data collection was conducted between June 2020 and December 2023. Qualitative data was audio recorded, transcribed verbatim, entered QSR NVIVO version 12 for management and coding.

### Data analysis

We used critical discourse analysis informed by the WPR approach as a theoretical framework to analyze and interpret the policy documents and interview data (43). First, policy documents were read in full and all sections that referred to human mobility and/or migration were inductively coded using the guiding questions. Attention was given to both the latent and the manifest meanings and contestations of these problem representations within the four Masterplans (43). Higher order themes were developed from the policy documents, the WPR analytic frame and reviews of the robustness of the reflected data. For the analysis of in-depth interviews, we used guidance provided by Bacchi and Bonham (56). We began by mapping precisely what was said in the interviews into codes based on the domains and constructs of the modified version of the Consolidated Framework for Implementation Research adapted for health equity and for use in low and middle income countries was used (54,55). This formed a baseline of normative practices in everyday implementation of NTD interventions as outlined in the Masterplans.

We then extracted specific segments from the interview data that made reference to human mobility and/or migration including a focus of the discursive practices that give rise to them (56).

Focusing on these extracted segments, we examined how the norms about implementation were evoked, which subjects were produced, objects created and the places that were produced as being legitimate (56). We identified the silences in providing migrant responsive care on four main dimensions; entitlement to health services, accessibility of health services, responsiveness of services to migrant needs and measures of achieving change due to implementation of equity measures (57). Based on this analysis we were able to map taken for granted realities and instances that were unusual or out of context (56). Specifically, this helped identify what policy actions were seen as possible or impossible and ultimately how they contributed either to the visibility or invisibility of migrant populations (58). Finally, we synthesized and reported the descriptions of problem representations, assumptions, silences and implications of these representations from the analyses.

### Ethical considerations

Ethical approval was obtained from ERES Converge Institutional Review Board (Ref. No. 2020-Jan-013) and the National Health Research Authority (Ref No: NHRA00012/4/03/2021). Study participants provided written consent prior to taking part in the study.

## RESULTS

### How is health equity represented in NTD Masterplans 2011-2026?

The Masterplans take a holistic view of health by incorporating dimensions of well-being and quality of life. Good health for all is recognized as a way of ensuring that every person is able to make a meaningful contribution to the development of the country. Various concepts that are commonly used to denote health equity were present in the Masterplans including *equity* and *socio-economic advantage*. Common terms used to refer to populations and groups facing inequities were *vulnerable populations, poor* and *marginalised, underserved populations, disadvantaged, socioeconomically disadvantaged* or *hard to reach*. The use of these terms could be associated with the development of an international health inequality consensus frame through the work of agencies such as the World Health Organisation which influences national level policy making (10). The constructions of equity in the four Masterplans were largely based on socioeconomic position. Population groups that were explicitly mentioned as facing inequities included young girls and women. Whereas socioeconomic status, lack of access to basic services such as water and hygiene, rural locations and gender were implicitly identified as positions that were associated with inequalities. This was described both generally and for specific diseases such as schistosomiasis.

> “Schistosomiasis affects the poorest communities and infections are particularly common among people living in rural or deprived urban or peri-urban settings. These populations typically have low socio-economic status with limited access to clean water and poor sanitation. Fishing, farming and collection of water fromsnail infested sources are the major risk factors for infection.” (Masterplan 2022-2026, P.26)

There was little mention of how these social factors contribute to differential vulnerability to NTDs or how the intersection of these axes of inequality result in unique challenges. None of the documents used population level quantitative or qualitative data as a basis for the existence of inequalities. The problematization of health equity in the Masterplans was closer to that of health inequalities than equity as differences between groups were stated to be present but not listed as being avoidable, preventable and unjust. Moreover, the recognition of the social determinants of health and their impact of NTD health outcomes grew in successive documents. Yet there was no mention of the structural determinants of health and health inequities. Explanatory accounts showing the relationship between the wider social, political and economic context and health inequities in the Masterplans were limited.

> “The health sector alone cannot address the multi-faceted determinants of NTDs, to this end the Ministry of health is working on collaborating with other relevant institutions including higher learning institutions in the control of NTDs using a One Health approach. It is with this realization that the control of NTDs requires moving beyond preventive chemotherapy to address the root social, ecological, environmental, animal and human factors.” (Masterplan 2022-2026, P.11)

The centrality of equity in the prevention, control and elimination of NTDs in Zambia was acknowledged in the different Masterplans often in the introductory sections. Importance was placed on the promotion of universal “*access to cost effective quality health care as close to the family as possible*”(Masterplan 2019-2023 P. xiii) including preventive chemotherapy, diagnosis, case management and reporting alongside protecting populations at risk from catastrophic out of pocket health expenditure due to NTDs within the strategic objectives of the different Masterplans. The Masterplan’s referenced other policy frameworks in Zambia such as the 7^th^ and 8^th^ National Development Plans and the National Health Strategic Plans, which have explicit objectives aimed at reducing poverty and vulnerability through a variety of services including *“healthcare and health related benefits for the poor and improved access to critical services”* (Masterplan 2022-2026 P.8). This framing of equity is aligned to the Sustainable Development Goals specifically; i. Target 3.3.5 which tracks progress being made in ensuring communities at risk of NTDs are able to receive the interventions that they need and ii. Target 3.8 which calls for the realization of universal health coverage (leaving no one behind) through actions geared towards preventing catastrophic financial expenditure and improving access to high quality services. It is also aligned to the push towards Primary health care that is aimed at bringing services as close to communities as possible. However, by looking at equity primarily through the lens of access of health services, the policy direction reflects downstream drift, where policy tools is oriented towards downstream rather than upstream determinants of health (10).

The Masterplans envision shared responsibility through multisectoral action as being key to addressing NTDs by bringing together Government Ministries, Private sector actors and actors beyond the health sector alongside communities living in NTD endemic areas. However, there is no guidance of how multisectoral collaboration will be operationalized in any of the plans limiting action. Community engagement and participation is highlighted as being essential in successful prevention and control but is largely centered on educational strategies. Furthermore, targeted strategies to promote community engagement from populations facing health inequities in relation to NTDs such as migrants are not highlighted in the policy documents Generally, there are very limited actions specifically targeted towards addressing health inequities including structural intervention. Consequently, there are no measures in place within the Masterplans to ascertain the progress being made to reduce the inequities faced by different population outside of treatment coverage and preventive chemotherapy coverage rates. This shows that health equity was not comprehensively problematized in the Masterplans.

### How are human mobility, migration or related terms represented in NTD Masterplans 2011-2026?

The impact of migration on NTDs was present in the Masterplans as evidenced by the recognition of different types of human mobility. Cross-border movement was commonly referenced in relation to either forced migrant (refugees) or voluntary migrants (foreign tourists). Whereas internal migration was either in relation to labour migration (fishing or farming) or environmental migration occasioned through climate change related disasters.

> “Displacement of people due to disasters leads to overcrowding that results into an enabling environment for the spread of diseases like trachoma and schistosomiasis. In addition, disasters and climate change can predispose people to newer infections from the places they have migrated to or they equally can introduce diseases to those areas. Lack of productivity among displaced persons increase their vulnerability which further increases their likelihood of poverty and thereby perpetuating the poverty cycle.” (Masterplan 2022-2026, P10)

Overall, the problematization of human mobility and migration in the Masterplans is poor. Across all four Masterplans, the construction of migration and its role in NTD prevention, control and treatment was scant indicating limited recognition of the role that migration plays on health. Where present, migration was framed as a risk factor for disease transmission, importation of new cases and a threat to disease control. However, there was no population level quantitative or qualitative data showing the magnitude of migration related inequities. The only interventions listed as being able to address migration related challenges were the establishment of cross border activities including development of cross border Memoranda of Understanding and surveillance to reduce the reintroduction of NTDs and to ensure continuity of care. However, facilitating cross border collaborations was largely seen as the mandate of the World Health Organisation. Migration is an issue that requires action across multiple sectors yet whole of system approaches were not listed as a potential solution to the inequalities.

### What assumptions underlie these representations of human mobility?

#### Territoriality associated with borders and internalization practices

Cross border mobility was problematized as being key to local transmission of NTDs through cross border importation of new diseases that are non-endemic in Zambia (onchocerciasis) or reintroduction of NTDs which have been eliminated (leprosy) by migrant groups. The Masterplans view borders as fixed orderings of space with cross border migration typically represented through refugees which excluded other types of cross border mobility and alluded to border fortification to prevent disease importation.

> “Mapping of Human Onchocerciasis will be particularly done in the border areas with countries endemic to the disease. The results of the mapping will be availed for use in the decision making on the disease together with the Ministry of Health.” (Masterplan 2022-2026, P.9-34)

> “The other challenge is influx of leprosy cases from countries bordering Zambia, especially among refugees. Greater investment into infrastructure and training are required for more accurate diagnosis and surveillance of leprosy in Zambia.” (Masterplan 2019-2023, P. 34)

This view of borders is different in communities, where community boundaries are more flexible. Implementers such as community health workers were seen to act as mediators using different strategies to improve service reach among mobile and migrant populations.

> “The only challenge I faced in the community was one client who had a field in Mozambique, but still I was to try by all means to meet him.” (Community Health Worker Interview 10, Luangwa District)

#### Medical nativism

Generally, migrants irrespective of mobility type, were seen in the Masterplans as sources of disease. This could be associated with historical portrayals of migrants as diseased others who introduce disease to native populations (59). This perception of migrants as disease carriers also brought to the fore differences in the levels of institutional visibility among migrant groups. Refugees for instance were more visible than other migrant groups. Such portrayals could affect societal views on migrants which can lead to discrimination, violence and xenophobia.

“This influx of migrants may cause a serious health risk in Zambia as they may import the infection from their home countries to the new settlements, causing a risk to other migrants. Some are exposed to NTD infection at the refugee settlements. The risk continues in the settlements which may have limited access to safe water and sanitation facilities.” (Masterplan 2022-2026, P.9-10)

#### Migrant deservingness/undeservingness

Irregular implementation of NTD interventions such as MDA or implementation during specific seasons and for short periods of time rendered labour migrants who have gone either to other districts or other countries incapable of accessing key health services.

“Because when you look at our setup people don’t usually come here to stay for longer periods, they will come and go so maybe those people you have talked to in the past three months others might shift and there is also a tendency of Zambians going to do cultivation to the other side of the river, so you find that when you go in the communities you find that certain individuals are not at their home place they are in Mozambique doing some cultivation and over a certain period they will still come back, so those interventions need to be regularly done so that at one time everyone will be aware of the focus and the direction that we are moving as an organization and as a district and other stakeholders.” (Key Informant Interview 2, Luangwa District)

> “The disadvantages we had the time that we were implementing that’s the time here people go for caterpillars, so you would find to say the time you will be in this catchment areas the whole community are going sleeping in the farms in the bush to go collect, so that usually used to give us challenges.” (Key Informant Interview 1, Nkeyema District)

#### Migrants impact universalization of health services

The Masterplans do not make reference to the right to health of some migrant and mobile populations such as refugees resulting in the absence of deliberate targeted strategies to enhance the reach of services. Limited access of NTD services was common in endemic districts and migration was felt to impact the realization of universal coverage goals for NTD interventions such as mass drug administration and improved water, sanitation and hygiene as it was difficult to plan for service delivery.

> “The other thing that is here people come from this is the food basket of western province, people come from all over Kalabo, Senanga they will come here specifically for that season. When they harvest they pack their maize they go back, meaning you will still have that data to say maybe the time that we are implementing the same MDA you go there, you follow it you just find, then they will come back when it’s time for farming, so I think those are the only barriers that I have seen, you can’t hope to say this village when I go there I will find them, you find to say they have shifted and since of the tobacco people keep shifting to virgin land whereby they start cutting trees again, so like the community keeps on moving, that’s the only challenge we have, they don’t have permanent home whereby everything is built according to the standard.” (Key Informant Interview 1, Nkeyema District)

Implementers’ efforts to provide services to migrant groups also brought out the view that inequalities arising from migration were not unidimensional. For instance, religious beliefs among cross-border migrants determined how willing they were to utilize health services such as medicine provided during MDA. These beliefs impacted their willingness to utilize health services.

“They were saying the bible says there will be different diseases towards the end of the world, so these diseases they are talking about are just fulfilling the prophecy of the bible and we do not need to take drugs, it is God who heals human beings. (Community Member Interview 5, Kapiri District)

Age was also identified as another aspect of social identity which alongside migration affected service use. This was particularly the case for school going children in schools that are locations different from their homes who could not take part in activities without parental consent.

> “The children go in schools, and we are passing at homes door by door, we may convince the parents at home, but the child at school will refuse because he believes the parents cannot allow that.” (Community Health Worker Interview 5, Kapiri District)

### What is left silent by the representations of human mobility and migration in NTD Masterplans?

We provide a brief summary of the key areas where the Masterplans are silent in relation to the promotion of migrant health specifically; entitlement to health services, accessibility of health services, responsiveness of services to migrant needs and measures of achieving change due to implementation of equity measures as illustrated in Table 2 (57).

**Table 1.**
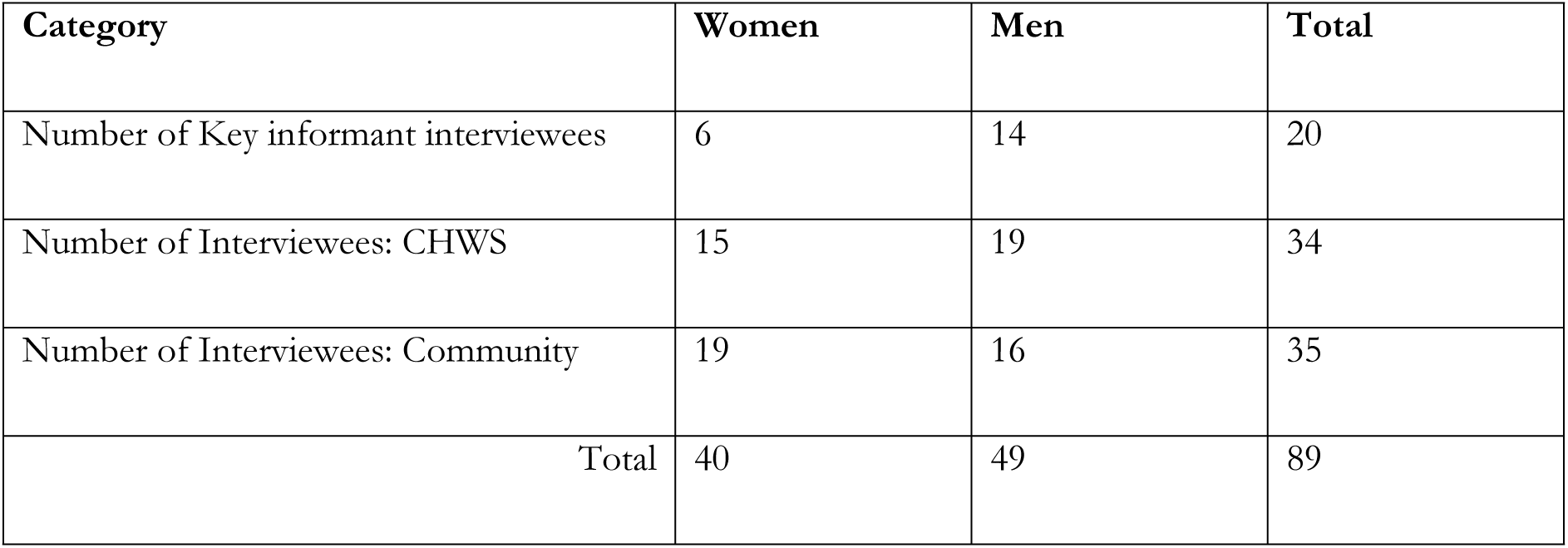
Summary of Study participants across the 4 study sites.

**Table 2.** Silences in problem representations of human mobility and migration in NTD Policy Discourse.

| Key areas | Silences |
| --- | --- |
| Entitlement of health services | • Masterplans do not articulate how the right to health will be realized in the face of different types of human mobility. |
| Equitable access to NTD Interventions | • Absence of provisions for interpretation services or translated materials in areas with high levels of internal and cross border mobility. |
| Migrant responsive health services | • Implementation plans fail to account for migration patterns<br>• Limited involvement of migrant communities and groups in service delivery.<br>• No requirement for training in health worker sensitivity to migrant health needs. |
| Measures of progress in reducing inequities | • Absence of population level data to highlight migration related inequalities.<br>• Limited involvement of migrant communities and groups.<br>• Migrant aware whole of system approaches missing. |

## DISCUSSION

Population mobility, encompassing all forms of movement (60), heightens migrant vulnerability to illness through disproportionate exposure to disease causing agents, barriers to accessing social services such as health care and the root causes of disease (61). Migration aware public policies are characterized by a sensitivity to population mobility, a recognition of the heterogeneity of migrant populations who pass through spaces of vulnerability that impact their health and well-being, they promote the use systems based strategies, are rooted in a health for all approach and are aware of mobility patterns at a regional level (5,13). Making health policies such as Neglected Tropical Disease Masterplans migration aware permits the integration of fairness and justice as a norm in policy processes ensuring that inequalities and social exclusion are addressed (62). Consequently improving health outcomes for migrant and mobile populations (63). Our study utilized a **policy as discourse** analysis to critique the problem representations of migration mobility and migration in NTD masterplans that have been developed in Zambia between 2011 and 2022. By identifying whether the masterplans underlying policy paradigms result in implementation gaps and inequalities due to the assumptions and problematization of population mobility (35).

We found that the current depictions of migration and population mobility within the different Neglected Tropical Disease Masterplans are not very comprehensive which hampers the type of strategies proposed to make services migrant and mobility responsive. Common representations of mobility through the lens of cross border migration and labour migration found in our analysis uphold a colonial idea of borders as instruments to keep people out (59). They ignore the dynamic nature of population mobility that has been existing in the country and the southern African regions for more than a hundred years. Thereby limiting the likelihood of the realization of the goals of free movement of people even in countries such as Zambia with open door policies (64). These representations also undervalue other forms of population mobility such as internal migration, undocumented migration, internal displacement or environmental related migration. For instance in Zambia, internal migration is more common than any other form of migration but has not been adequately addressed in health policy responses including the Masterplans (65). Manji’s review of government policies and legislation in South Africa found that very few engage with migration and population mobility with migration often being depicted as problematic and recognizing very few forms of mobility (18). Even where policies that recognize population mobility exist they may be limited in scope to a few health services and diseases such as HIV, TB or COVID, cater for physical health and not mental health or tailored to ignoring to a small group of migrants (8).

Additionally, protocols and guidelines used within health facilities do not often include provisions for migrants even where laws have mandated the creation of migrant aware services (66). Training of frontline workers using standards such as the WHO Refugee and Migrant Health Competency Standards for Health workers can ensure that they are able to provide migrant responsive NTD services and health services in general. This can be done both in service and among students. As it equips providers with the capacity to understand and address the health and social needs of different migrant populations (67).

The rights of migrants to the highest possible standard of physical and mental health is reliant on the development, implementation and monitoring of migration governance responses (65). Mobile and migrant populations tend to receive integrated health services provided across different organizations found in one or more countries with varying missions, capacities and commitments to furthering migrant health. Local responses from actors working in border regions have exemplified how the health sector can be instrumental in the development of migration aware approaches. As they require health providers to work with a broad range of service providers beyond the health care system in order to strike a balance between navigating migration related challenges associated with freedom of movement alongside challenges related to access to health.

Though multisectoral action has been listed as being critical for the addressing the structural determinants of migrant health inequities, existing policies including the NTD Masterplans do not provide operational plans on how this can be realized in practice. Intersectoral fora such as district and provincial level migration and health taskforces, which bring together different civil society, multilateral organizations, state structures and district health services have been shown to improve access to health services for cross border migrant and can be applied for other forms of migration (5). This is because such cross system interventions align services across different systems which enhances coordination and reduce fragmentation of care (68). They have also been shown to be effective in improving service and health outcomes in various sectors including child welfare (68). Potential strategies that can be selected will include; administrative strategies (Staff co-location, joint funding, collaborative structures), Frontline practices (data sharing) and third party facilitation (brokering, resource support and regional coordination) (69).

Equity relevant data is necessary if the goals of equitable policy making are to be realized (71). According to Slootjes, policies that aimed at addressing inequalities faced by vulnerable populations such as migrants, often lack targets and goals through which progress being made to address disparities are measured for increased accountability (Slootjes, 2021). Detailed data on health outcomes across different migration groups is often absent or threadbare which renders migrants invisible from health information systems (Vidal and Wickramage, 2023). One of the key gaps identified in the Zambia’s National Migration Policy 2022 is the absence of adequate data that can be used to inform better migration governance and management on aspects such as mobility patterns, migrant profiles and migrant health needs. This is evident in the masterplans which do not use any qualitative or quantitative data to depict the impact of migration on NTD service utilisation and health outcomes. The absence of data on local migration dynamics means that strategies being proposed at national, provincial and district levels are not aligned with local population mobility patterns. Further, accountability tools that ensure that commitments made to address migrant health inequities are met, are needed so that these intentions are realized (71).

Currently, even when interventions are put in place to address migrant related inequalities, the absence of evaluative systems hampers the capacity to measure the impact of these measures do to the absence of robust data capture systems. Given the paucity of indicators, policy actors can collaboratively identify standardized data elements that can be used to measure and monitor migrant health while at the same time advocating for enhanced data collection methods and data sharing to improve service delivery and evidence informed decision making for migrant and mobile populations (67). Methods such as Population Mobility Mapping and Events based Surveillance can be used to characterize mobility dynamics and generate an evidence base to inform decision making (70,72).

Advocacy efforts can be useful in raising awareness on the need for migration aware approaches and promote policy coherence across different sectors involved in the provision of social services to migrants (70). It can also be used to raise awareness on the legal entitlements of migrants when it comes to accessing health services and counter public perceptions on migrant deservingness of social services such as health. Our analysis found limited engagement of migrant and mobile populations in migration governance. The provision of opportunities through which mobile and migrant populations could be actively involved in decision making would promote the development and implementation of services and policies that are migrant responsive. It would also empower this populations to develop the capacities through which they can collectively advocate for their health. For instance, intersectoral action is critical in promoting migrant health but it ought to go beyond a focus on individual level factors (such as access) to explicit focus on upstream conditions (73).

## Conclusion

Public policies play a critical role in ensuring health systems can be made more responsive to the needs of cross border and internal migrants. Our current study explored the problem representations of mobility and migration within NTD Masterplans, how these representation arose and what is excluded as a result. Though the mobility is poorly problematized in the Masterplans, we identified different ways through policy design and implementation can strengthened such as using more comprehensive and standardized definitions of migration and migrant groups in public policies, capturing migration data and using it to inform decision making as well as establishing accountability mechanisms for policy actions.

## Data Availability

All relevant data are within the paper and its Supporting Information files.

## REFERENCES

1. Spitzer DL. Intersectional and embodied: migration as a social determinant of health. In: Handbook on the Social Determinants of Health. Edward Elgar Publishing; 2025. p. 204–19.

2. Recchi E, Safi M. Introduction: human mobility as hallmark of our age. In: Handbook of Human Mobility and Migration. Edward Elgar Publishing; 2024. p. xii–xxvi.

3. Abubakar I, Aldridge RW, Devakumar D, Orcutt M, Burns R, Barreto ML, et al. The UCL– Lancet Commission on Migration and Health: the health of a world on the move. The Lancet. 2018;392(10164):2606–54.

4. Castañeda H. Migration and Health: Critical Perspectives. Taylor & Francis; 2022.

5. Vearey J, Modisenyane M, Hunter-Adams J. Towards a migration-aware health system in South Africa: a strategic opportunity to address health inequity. South African health review. 2017;2017(1):89–98.

6. McAuliffe M, Oucho LA. World Migration Report 2024. Geneva: International Organisation for Migration (IOM); 2024.

7. International Organization for Migration (IOM). Africa Migration Report (Second edition). Connecting the threads: Linking policy, practice and the welfare of the African migrant. Addis Ababa: IOM; 2024.

8. Bojorquez I, Cubillos-Novella A, Arroyo-Laguna J, Martinez-Juarez LA, Sedas AC, Franco-Suarez O, et al. The response of health systems to the needs of migrants and refugees in the COVID-19 pandemic: a comparative case study between Mexico, Colombia and Peru. The Lancet Regional Health–Americas. 2024;

9. World Health Organization. Global research agenda on health, migration and displacement: strengthening research and translating research priorities into policy and practice. Global research agenda on health, migration and displacement: strengthening research and translating research priorities into policy and practice. 2023;

10. Lynch J. Regimes of inequality: the political economy of health and wealth. Cambridge University Press; 2020.

11. Anand S. Measuring disparities in health: methods and indicators. Challenging Inequalities in Health: From Ethics to Action. T. Evens, M. Whitehead, F. Diderichsen, A. Bhuiya and M. Wirth. 2001;

12. Bambra C, Lynch J, Smith KE. Introduction: The holy grail of reducing health inequalities. In: Getting Better. Policy Press; 2025. p. 1–14.

13. Vearey J. Healthy migration: a public health and development imperative for South (ern) Africa: forum-opinion. South African Medical Journal. 2014;104(10):663–4.

14. Goldenberg SM, Fischer F. Migration and health research: past, present, and future. BMC Public Health. 2023;23(1):1425.

15. Cf O. Transforming our world: the 2030 Agenda for Sustainable Development. United Nations, New York. 2015;

16. Mosca DT, Vearey J, Orcutt M, Zwi AB. Universal Health Coverage: ensuring migrants and migration are included. Global Social Policy. 2020;20(2):247–53.

17. Wickramage K, Vearey J, Zwi AB, Robinson C, Knipper M. Migration and health: a global public health research priority. BMC public health. 2018;18:1–9.

18. Manji K, Perera S, Hanefeld J, Vearey J, Olivier J, Gilson L, et al. An analysis of migration and implications for health in government policy of South Africa. International journal for equity in health. 2023;22(1):82.

19. Onarheim KH, Rached DH. Searching for accountability: can the WHO global action plan for refugees and migrants deliver? BMJ Global Health. 2020;5(6):e002095.

20. Avaria A, Ventura-Garcia L, Sanmartino M, Van der Laat C. Population movements, borders, and Chagas disease. Memórias do Instituto Oswaldo Cruz. 2022;117:e210151.

21. Maritim P, Chewe M, Munakaampe MN, Silumbwe A, Sichone G, Zulu JM. Applying community health systems lenses to identify determinants of access to surgery among mobile & migrant populations with hydrocele in Zambia: A mixed methods assessment. PLOS Global Public Health. 2023;3(7):e0002145.

22. Brady MA, Toubali E, Baker M, Long E, Worrell C, Ramaiah K, et al. Persons ‘never treated’in mass drug administration for lymphatic filariasis: identifying programmatic and research needs from a series of research review meetings 2020–2021. International health. 2023;ihad091.

23. Hopkins AD. Neglected tropical diseases in Africa: a new paradigm. International Health [Internet]. 2016 Mar 1 [cited 2021 Apr 17];8(suppl_1):i28–33. Available from: 10.1093/inthealth/ihv077

24. Expanded Special Project for Elimination of Neglected Tropical Diseases. Country NTD Master Plan 2021-2025: Framework for Development.

25. Powell BJ, Waltz TJ, Chinman MJ, Damschroder LJ, Smith JL, Matthieu MM, et al. A refined compilation of implementation strategies: results from the Expert Recommendations for Implementing Change (ERIC) project. Implementation Science. 2015;10(1):1–14.

26. World Health Organization. Integrating neglected tropical diseases into global health and development: fourth WHO report on neglected tropical diseases. World Health Organization; 2017.

27. Gaias LM, Arnold KT, Liu FF, Pullmann MD, Duong MT, Lyon AR. Adapting strategies to promote implementation reach and equity (ASPIRE) in school mental health services. Psychology in the Schools. 2021;

28. Unitinng to Combat Neglected Tropical Diseases. Translating the London Declaration into Action: Meeting Report. 2012 Nov.

29. Mbabazi P, Del Pino S, Ducker C, Dean L, Broekkamp H, Prasetyanti W, et al. Promoting gender, equity, human rights and ethnic equality in neglected tropical disease programmes. Transactions of The Royal Society of Tropical Medicine and Hygiene. 2021;115(2):188–9.

30. Christine Masong M, Ozano K, Tagne MS, Tchoffo MN, Ngang S, Thomson R, et al. Achieving equity in UHC interventions: who is left behind by neglected tropical disease programmes in Cameroon? Glob Health Action [Internet]. 2021;14(1):1886457. Available from: https://www.embase.com/search/results?subaction=viewrecord&id=L634437256&from=export

31. Bambra C, Fox D, Scott-Samuel A. Towards a politics of health. Health promotion international. 2005;20(2):187–93.

32. Fischer F, Torgerson D, Durnová A, Orsini M. Introduction to critical policy studies. In: Handbook of critical policy studies. Edward Elgar Publishing; 2015. p. 1–24.

33. Duncan S, Reutter L. A critical policy analysis of an emerging agenda for home care in one Canadian province. Health & Social Care in the Community. 2006;14(3):242–53.

34. Came H, O’Sullivan D, McCreanor T. Introducing critical Tiriti policy analysis through a retrospective review of the New Zealand Primary Health Care Strategy. Ethnicities. 2020;20(3):434–56.

35. Jessop B, Sum NL. The cultural political economy approach to public policy. In: Handbook on Critical Political Economy and Public Policy. Edward Elgar Publishing; 2023. p. 36–48.

36. Donger E, Leigh J, Fuller A, Leaning J. Refugee youth in Lusaka: A comprehensive evaluation of health and wellbeing. UNHCR and Harvard FXB. 2017;1–17.

37. International Organisation for Migration. Migration in Zambia: A country profile 2019. 2019.

38. Zambia Statistics Agency, Ministry of Labour and Social Security. Zambia Labour Migration Report 2022. 2024.

39. Zulu JM, Maritim P, Silumbwe A, Halwiindi H, Mubita P, Sichone G, et al. Unlocking trust in community health systems: lessons from the lymphatic filariasis morbidity management and disability prevention pilot project in Luangwa District, Zambia. International Journal of Health Policy and Management. 2022;11(Special Issue on CHS-Connect):80–9.

40. UNHCR. Zambia: Analysis of the gaps in the protection of Refugees. Zambia; 2007 Sep.

41. Howarth D, Griggs S. Poststructuralist discourse theory and critical policy studies: interests, identities and policy change. In: Handbook of critical policy studies. Edward Elgar Publishing; 2015. p. 111–27.

42. Shaw SE. Reaching the parts that other theories and methods can’t reach: How and why a policy-as-discourse approach can inform health-related policy. Health: 2010;14(2):196–212.

43. Bacchi C, Goodwin S. Poststructural policy analysis: A guide to practice. Springer; 2016.

44. Schneider E, Brad A, Brand U, Krams M, Lenikus V. Historical-materialist policy analysis of climate change policies. In: Handbook on Critical Political Economy and Public Policy. Edward Elgar Publishing; 2023. p. 110–26.

45. Bacchi C. Policy as discourse: What does it mean? Where does it get us? Discourse: studies in the cultural politics of education. 2000;21(1):45–57.

46. Chung A, Zorbas C, Peeters A, Backholer K, Browne J. A critical analysis of representations of inequalities in childhood obesity in Australian health policy documents. International Journal of Health Policy and Management. 2022;11(9):1767.

47. Ministry of Health. Zambia Neglected Tropical Diseases Masterplan 2022-2026. 2023.

48. Matapo BB, Mpabalwani EM, Kaonga P, Simuunza MC, Bakyaita N, Masaninga F, et al. Lymphatic Filariasis Elimination Status: Wuchereria bancrofti Infections in Human Populations After Five Effective Rounds of Mass Drug Administration in Zambia. 2023

49. Nsakashalo-Senkwe M, Mwase E, Chizema-Kawesha E, Mukonka V, Songolo P, Masaninga F, et al. Significant decline in lymphatic filariasis associated with nationwide scale-up of insecticide-treated nets in Zambia. Parasite Epidemiol Control [Internet]. 2017;2(4):7–14. Available from: https://www.embase.com/search/results?subaction=viewrecord&id=L618310923&from=export

50. Mwale C, Mumbi W, Funjika M, Sokesi T, Silumesii A, Mulenga M, et al. Prevalence of Trachoma in 47 Administrative Districts of Zambia: Results of 32 Population-Based Prevalence Surveys. Ophthalmic Epidemiol. 2018 Dec;25(sup1):171–80.

51. World Health Organization. Accelerating work to overcome the global impact of neglected tropical diseases: a roadmap for implementation. World Health Organization; 2012.

52. World Health Organization. Ending the neglect to attain the sustainable development goals: a road map for neglected tropical diseases 2021–2030. World Health Organization; 2020.

53. Burgess RA. Rethinking global health: Frameworks of power. Taylor & Francis; 2024.

54. Means AR, Kemp CG, Gwayi-Chore MC, Gimbel S, Soi C, Sherr K, et al. Evaluating and optimizing the consolidated framework for implementation research (CFIR) for use in low-and middle-income countries: a systematic review. Implementation Science. 2020;15(1):1–19.

55. Woodward EN, Singh RS, Ndebele-Ngwenya P, Castillo AM, Dickson KS, Kirchner JE. A more practical guide to incorporating health equity domains in implementation determinant frameworks. Implementation science communications. 2021;2(1):1–16.

56. Bacchi C, Bonham J. Poststructural interview analysis: Politicizing“personhood”. In Palgrave Macmillan; 2016.

57. Ingleby C, Ayalew M, Ponde T. Promoting human rights and access to health services in prisons in Southern Africa: VSO, UNODC and SDC working together to reduce HIV and improve the health of incarcerated populations. J Int AIDS Soc [Internet]. 2016;19((Ingleby C.) Programme Development and Policy Team, Voluntary Service Overseas, London, United Kingdom):173. Available from: https://www.embase.com/search/results?subaction=viewrecord&id=L615728454&from=e xport

58. Özdemir GŞ. Intersectional (In) visibility: experiences of irregular migrants in Barcelona. Journal of Ethnic and Migration Studies. 2024;50(8):1862–85.

59. Castaneda H. Migration and Health. In: Routledge International Handbook of Critical Issues in Health and Illness. Routledge; 2021. p. 276–88.

60. Forray AI, Oltean O, Hanft-Robert S, Madzamba R, Liem A, Schouten B, et al. Uncovering multi-level mental healthcare barriers for migrants: a qualitative analysis across China, Germany, Netherlands, Romania, and South Africa. BMC Public Health. 2024;24(1):1593.

61. Slootjes J. Healing the Gap: Building Inclusive Public-health and Migrant Integration Systems in Europe. Brussels: Migration Policy Institute Europe[Google Scholar]. 2021;

62. Fainstein SS. Social justice and urban policy deliberation: balancing the discourses of democracy, diversity and equity. In: Handbook of critical policy studies. Edward Elgar Publishing; 2015. p. 190–204.

63. Gustavson AM, Vincenzo J, Miller MJ, Falvey JR, Lee JL, Fashaw-Walters S, et al. Equitable implementation of innovations to promote successful aging in place. Journal of the American Geriatrics Society. 2022;71(2):683.

64. Ndlovu-Gatsheni SJ. Decolonising borders, decriminalising migration and rethinking citizenship. Crisis, identity and migration in post-colonial Southern Africa. 2018;23–37.

65. Vearey J, Orcutt M, Gostin L, Braham CA, Duigan P. Building alliances for the global governance of migration and health. bmj. 2019;366.

66. Aith FMA, Forsyth C, Shikanai-Yasuda MA. Chagas disease and healthcare rights in the Bolivian immigrant community of São Paulo, Brazil. Tropical Medicine and Infectious Disease. 2020;5(2):62.

67. Africa Centre for Migration & Society. It is just because I am a foreigner: Making sexual and reproductive health and rights a reality for migrants, refugees and asylum seekers in South Africa. 2023 Mar.

68. Bunger AC, Chuang E, Girth A, Lancaster KE, Gadel F, Himmeger M, et al. Establishing cross-systems collaborations for implementation: protocol for a longitudinal mixed methods study. Implementation Science. 2020;15:1–14.

69. Chuang E, Bunger A, Smith R, Girth A, Phillips R, Miech E, et al. Collaboration strategies affecting implementation of a cross-systems intervention for child welfare and substance use treatment: a mixed methods analysis. Implementation Science Communications. 2024;5(1):127.

70. International Organization for Migration. Health, Border and Mobility Management Framework. Geneva.; 2021.

71. Schultz S, Zorbas C, Peeters A, Yoong S, Backholer K. Strengthening local government policies to address health inequities: perspectives from Australian local government stakeholders. International Journal for Equity in Health. 2023;22(1):119.

72. Chotun N, Eaton J, Anagbogu IA, Tesfahunei HA, Shawa S, Karutu C, et al. Sustaining success through strategies for post-elimination management of neglected tropical diseases in African Union Member States. Frontiers in Tropical Diseases. 2024;5:1421522.

73. Fisher M, Baum FE, MacDougall C, Newman L, McDermott D, Phillips C. Intersectoral action on SDH and equity in Australian health policy. Health Promotion International. 2017;32(6):953–63.

